# Emergence and Spread of the SARS-CoV-2 Omicron Variant in Alberta Communities Revealed by Wastewater Monitoring

**DOI:** 10.1101/2022.03.07.22272055

**Authors:** Casey R.J. Hubert, Nicole Acosta, Barbara J. Waddell, Maria E. Hasing, Yuanyuan Qiu, Meghan Fuzzen, Nathanael B.J. Harper, María A. Bautista, Tiejun Gao, Chloe Papparis, Jenn Van Doorn, Kristine Du, Kevin Xiang, Leslie Chan, Laura Vivas, Puja Pradhan, Janine McCalder, Kashtin Low, Whitney E. England, Darina Kuzma, John Conly, M. Cathryn Ryan, Gopal Achari, Jia Hu, Jason L. Cabaj, Chris Sikora, Larry Svenson, Nathan Zelyas, Mark Servos, Jon Meddings, Steve E. Hrudey, Kevin Frankowski, Michael D. Parkins, Xiaoli (Lilly) Pang, Bonita E. Lee

## Abstract

Wastewater monitoring of SARS-CoV-2 allows for early detection and monitoring of COVID-19 burden in communities and can track specific variants of concern. Targeted assays enabled relative proportions of SARS-CoV-2 Omicron and Delta variants to be determined across 30 municipalities covering >75% of the province of Alberta (pop. 4.5M) in Canada, from November 2021 to January 2022. Larger cities like Calgary and Edmonton exhibited a more rapid emergence of Omicron relative to smaller and more remote municipalities. Notable exceptions were Banff, a small international resort town, and Fort McMurray, a more remote northern city with a large fly-in worker population. The integrated wastewater signal revealed that the Omicron variant represented close to 100% of SARS-CoV-2 burden prior to the observed increase in newly diagnosed clinical cases throughout Alberta, which peaked two weeks later. These findings demonstrate that wastewater monitoring offers early and reliable population-level results for establishing the extent and spread of emerging pathogens including SARS-CoV-2 variants.

## Introduction

The COVID-19 pandemic has led to rapid scientific progress in wastewater-based surveillance of community infections. Measuring levels of RNA from SARS-CoV-2 in sewage samples began being used as a complementary surveillance tool early in the pandemic, resulting in hundreds of wastewater COVID-19 monitoring groups and online dashboards around the world (Naughton et al. 2021), including in Alberta (https://covid-tracker.chi-csm.ca). This strategy is premised on the fecal shedding of SARS-CoV-2 by infected individuals (Cevik et al. 2021; Yuan et al. 2021) and enabled by modifying RT-qPCR workflows used for diagnosing patients to quantify viral RNA in sewage sampled at wastewater treatment plants (WWTPs) or other nodes within the sewer network (Acosta et al. 2021a, b; Qiu et al. 2022) at regular intervals. Teams in Alberta and elsewhere demonstrated during pandemic waves that wastewater is a leading indicator of COVID-19, with results typically preceding clinical diagnosed cases by 4-6 days (e.g., D’Aoust et al. 2021, Medema et al. 2020; Nemudryi et al. 2020; Randazzo et al. 2020). Sampling, testing and rapidly reporting wastewater virus RNA levels provides early warning of the population-wide disease burden to policy makers, health officials and the public, enabling evidence-based decision making for preparedness and disease control.

On November 24, 2021, South Africa first reported the emergence of a novel SARS-CoV-2 variant associated with rapid community transmission in the Gauteng province (WHO, 2021). By November 26^th^ the World Health Organization had labelled Omicron as a new variant of concern (VOC). Omicron was subsequently rapidly identified in countries around the world, including in Canada where on November 28^th^ cases were detected in inbound international travellers. Alberta’s first case of Omicron from clinical specimen testing was confirmed on November 30. By December and into January this virus had spread rapidly throughout large and smaller communities, prompting re-introduction of public health restrictions.

Wastewater testing can also differentiate changes in disease burden caused by different VOCs in communities (Lee et al. 2021). As soon as viral genomes of VOCs become available within the international scientific community, e.g., via GISAID (Elbe & Buckland-Merrett, 2017), variant-specific PCR primers and probes can be developed and deployed on regularly collected wastewater samples to understand the dynamics of community disease burden caused by VOCs (Peterson et al. 2022). While sequencing viral genomes from wastewater is technically feasible, either via targeted amplicon tiling protocols (Rios et al. 2021; Lin et al. 2021) or shotgun metagenomics (Rothman et al. 2021; Pérez-Cataluña et al. 2022), these comprehensive approaches are significantly more costly and time consuming than targeted RT-qPCR screening of RNA extracted from wastewater that can provide accurate data on VOCs at a fraction of the cost, and in near-real time. In Alberta wastewater is sampled, processed and analyzed in university laboratories in Calgary and Edmonton, and reported to health officials and online to the public two days after sampling. In this study, variant-specific PCR assays were employed to assess the emergence and temporal change in prevalence of the Omicron and Delta variants in Alberta by monitoring wastewater in 30 municipalities ranging from small towns (pop. <10,000) to large cities (pop. >1M) up to three times per week. Wastewater surveillance demonstrated changes in COVID-19 burden associated with emergence of the new Omicron variant for >75% of Alberta’s population of 4.5M between late November 2021 and mid-January 2022.

## Methods

Wastewater was collected from municipal WWTPs across the province as 24-h composite samples up to 3 times per week. RNA was isolated from wastewater using either affinity binding columns that purify nucleic acids directly (Whitney et al. 2021) or ultrafiltration followed by RNA extraction (Qiu et al. 2022). These two approaches were used to process 233 and 209 WWTP samples, respectively (Figure 1), with the same method applied to a given sampling site over time throughout the entire study period. Wastewater samples from three geographically disparate WWTPs in Calgary, Fort McMurray and Lethbridge, comprising 11% of all samples in the study, were processed using both methods for comparison and revealed no difference (Mann-Whitney test P = 0.46, 0.39 and 0.59, respectively; Fig. S1).

**Figure 1.**
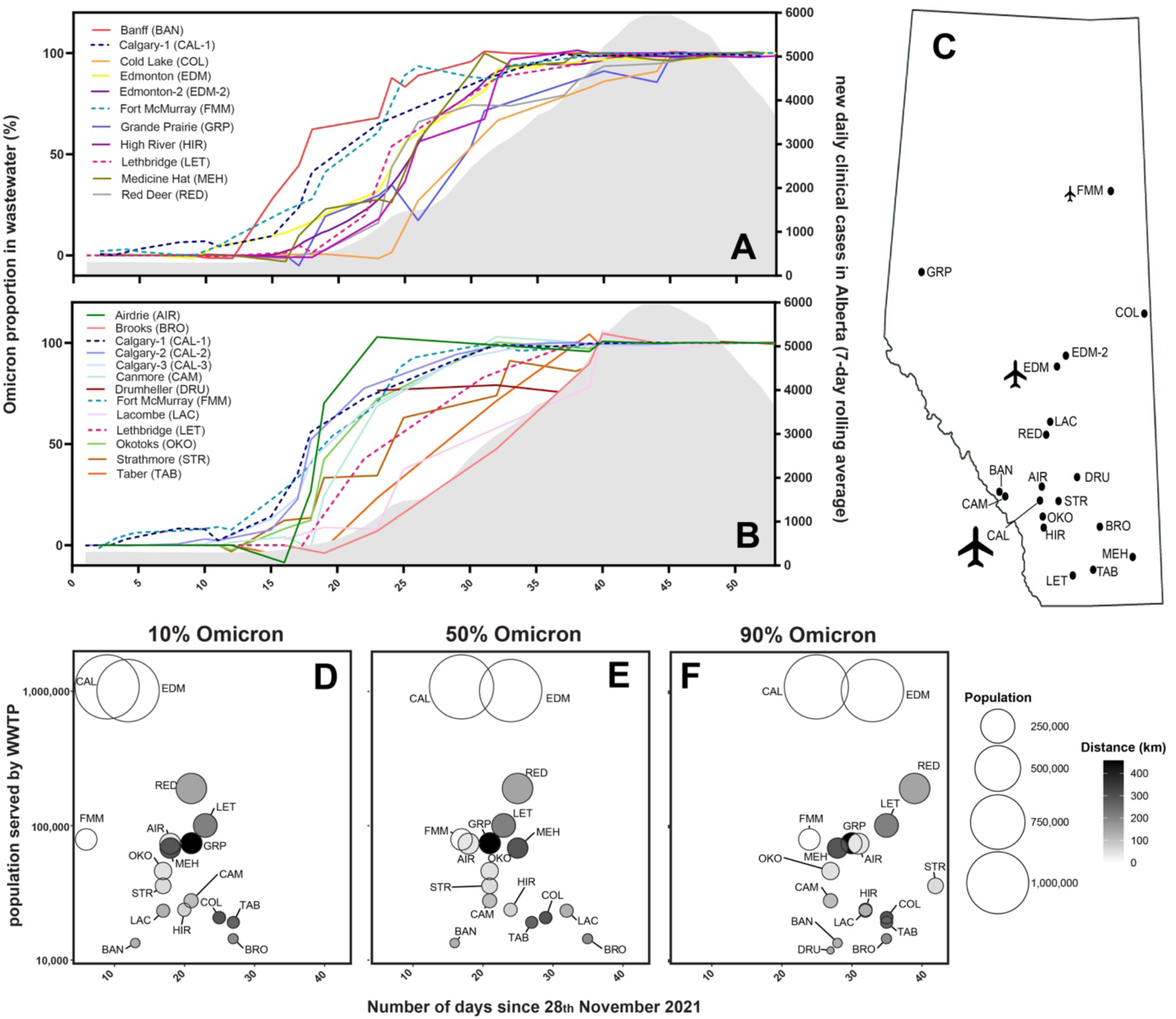
Proportion of Omicron relative to Delta SARS-CoV-2 variants in community wastewater samples assessed using RT-qPCR assays for specific variants following sample processing using ultrafiltration (A) or affinity columns (B). Lines of best fit plotted with second order smoothing are shown for different WWTPs, including 3 that had samples processed using both ultrafiltration (A) and affinity columns (B) for comparison (see dashed lines for Calgary-1, Fort McMurray and Lethbridge; Fig. S1). Monitoring lasted for 53 days beginning on November 28^th^ (plotted as consecutive days on the x-axes). The grey shaded area on the right side (A, B) shows the 7-day rolling average of new clinical cases reported in Alberta (right y-axis), which increased after the Omicron variant was predominant in municipal wastewater from 30 communities sampled at 21 WWTPs throughout the province (C). Calgary and Edmonton are served by 3 and 2 WWTPs, respectively (A, B), and some individual WWTPs also serve several municipalities (e.g., Edmonton-2 serves 6 others; Red Deer serves 3 others; Calgary’s WWTPs serve 3 others). The timing (in days) of the Omicron-to-Delta ratio passing 10%, 50% and 90% of community COVID-19 burden (D-F) reveals general trends of decreasing population size (bubble diameter and y-axis) and distance from the nearest airport in Calgary, Edmonton or Fort McMurray (bubble shading). Bubble plots only include data from Calgary-1 and Edmonton-1 WWTPs (the largest WWTP from each city), scaled to the population of the corresponding sewershed sub-catchment in those cities.

RNA quantification by RT-qPCR incorporated a newly designed set of assays that selectively amplify the B.1.1.529 Omicron variant or the Delta variant by targeting mutations in amino acids 203 and 204 of the nucleocapsid gene. Omicron and Delta were the only two variants detected in Alberta by clinical screening during the study period (Alberta Health, 2022). Total SARS-CoV-2 levels were quantified with widely used universal assays targeting the N1 & N2 regions of the nucleocapsid gene in the wild type virus (Acosta et al. 2021a, b) and all other VOCs identified to date. Omicron, Delta and total SARS-CoV-2 assays were triplexed together enabling an Omicron-to-Delta ratio to be estimated in each wastewater sample using the Omicron signal (R203K-G204R assay) and the Delta signal (R203M assay). This allowed the emergence and prevalence of Omicron to be tracked at the population level throughout the province.

The daily number of new cases of COVID-19 clinically diagnosed across the province were collected from Data Analytics of Alberta Health Services, and are reported using a 7-day rolling average.

## Results

Wastewater separation, identification and quantification of SARS-CoV-2 is intrinsically more complicated than conducting the same PCR strategy on clinical samples (i.e., nasopharyngeal swabs). It is not normally recommended that results be directly compared between different WWTPs, due to intrinsic heterogeneities, e.g., physiochemical differences manifesting different PCR inhibition potential, different proportions of urban, industrial and agricultural inputs, and different flow rates and distances impacting signal degradation (Pecson et al. 2021). While these limitations apply to total SARS-CoV-2 quantification, they are mitigated when determining Omicron-to-Delta ratios within the same multiplex RT-qPCR reaction, since RNA genomes derived from either variant are expected to react similarly to the factors mentioned above.

Figure 1 shows the emergence of the Omicron variant (corresponding to the displacement of the Delta variant) in different Alberta communities. Omicron was first detected in Alberta community wastewater during late November and early December (Fig. 1; Table S1). In Calgary, four consecutive samples collected during December 5-9 revealed the sustained presence of 3-9% Omicron (compared to >90% Delta) among infected individuals contributing to the sewershed in this cosmopolitan city of 1.3 million people. Omicron was first detected in wastewater in the capital city of Edmonton (pop. 1.1 million) on December 10^th^ (15% Omicron; 85% Delta). The rate of increase of Omicron in the international resort town of Banff was higher than in larger cities such as Calgary and Edmonton (Fig. 1) and surpassed 80% in thrice weekly samples taken during December 20-23. By this time, Calgary and Edmonton had just surpassed 50%, and the proportion of Omicron infections was growing in smaller bedroom communities adjacent to these two large urban centres (e.g., Okotoks, High River, Strathmore and especially Airdrie, which are all <70 km away from Calgary; Fig. 1, Fig. S2). Communities that experienced the most delayed emergence of Omicron were smaller and more remote, with Brooks (pop. 14,451; 190 km from Calgary) and Taber (pop. 19,070; 263 km from Calgary) not reaching high proportions of Omicron until December 29^th^.

## Interpretation

The expected general trend demonstrated by this analysis of objective wastewater evidence is that large cities encounter the emergence of a new virus before the smaller centres farther away from the cities, but with notable exceptions. Banff is 127 km west of Calgary and experienced a more rapid onset of Omicron infection than anywhere else in the province despite the resident population in Banff (pop. 13,427) being <1% of Calgary. Banff is an international resort community in Banff National Park – Canada’s busiest national park – which attracts >4 million visitors annually from around the world (Banff, 2022). Early detection of Omicron in Banff may correspond to attracting tourists at the onset of the ski season in November and December. Interestingly the nearby and slightly larger mountain town of Canmore (pop. 27,664) located 105 km east of Calgary (22 km east of Banff, and outside the national park), experienced a much later onset of Omicron infection. This is likely related to Canmore hosting fewer international tourists than Banff, and featuring much less high-density dormitory-style living among the worker population in Banff that supports the tourism industry.

More remote communities located a greater distance away from Alberta’s large international airports exhibited later emergence of the Omicron variant (Fig. 1; Fig. S2). The Calgary International Airport serves 16 million travellers per year with direct flights arriving from 15 countries (YYC, 2022), compared to 8 million travellers and 6 international connections for the Edmonton International Airport (YEG, 2022). Plotting Omicron dynamics in Alberta municipalities as a function of distance from Calgary (Fig. S2) suggests a link to international travel. A notable exception to this trend is Omicron emergence in Fort McMurray. Despite being a remote, relatively small (pop. 79,205) northern city that is farther from Calgary than any other municipality sampled, Fort McMurray exhibited an Omicron emergence comparable to Calgary’s rapid onset. Fort McMurray has one of Canada’s busiest airports to accommodate shift workers commuting from across Canada to work in the oil sands industry (YMM, 2022). This high level of contact with other parts of the country is likely to facilitate rapid introduction of an emerging virus such as the Omicron variant. Workers travelling to Fort McMurray from other provinces, or Alberta’s major urban centres, likely contributed to accelerated Omicron emergence relative to other smaller and/or remotely situated Alberta municipalities.

Wastewater results also demonstrate that the emergence of Omicron was the driver of clinical cases increasing in December and January during Alberta’s 5^th^ wave (grey shaded area in Fig. 1A, B). During this time COVID-19 public health surveillance shifted to focus PCR testing on patients at risk for severe illness and eligible for early treatment, patients presenting to emergency wards with more serious illness, and essential workers (AHS, 2022). This resulted in PCR testing dramatically underestimating total disease burden in the population as a whole relative to earlier waves. Clinical cases that were reported still show a steep increase after the emergence and propagation of Omicron revealed by wastewater testing. These dynamics mirror Alberta’s shift from Delta to Omicron infections confirmed by screening sub-sets of clinical samples using PCR assays for VOCs and associated viral genome sequencing, which revealed Omicron to be >50% by December 16^th^ and >95% by December 28^th^ (Alberta Health, 2022). This demonstrates that wastewater surveillance reliably provides important information needed and acted upon by public health officials.

VOC information derived from viral genome sequencing of clinical samples is typically non-random, e.g, being intentionally biased towards clinical cases of interest (e.g., outbreaks; hospitalizations) or incoming international travellers (e.g., Williams et al. 2021). Similarly, clinical PCR testing is susceptible to changes in testing policies, capacity limitations or individuals not getting tested, e.g., by personal choice or when infections are asymptomatic (Green et al. 2021). Wastewater testing on the other hand offers an unbiased representation of disease prevalence, capturing all individuals and groups contributing to the sewershed for a tiny fraction of the cost of clinical testing on a per-capita basis. In large cities like Calgary and Edmonton with over one million residents (Fig. 1D-F; Fig. S1) monitoring wastewater for COVID-19 community burden costs only a few cents per person per year (based on three times weekly testing in Alberta) and can provide objective information about community infection to public health authorities, policy makers and the public in near real time. With COVID-19 clinical testing strategies and resources become more targeted, wastewater testing offers an important, early, objective population-based metric of disease burden surveillance that can be easily adapted for emerging pathogens throughout large jurisdictions like Alberta.

## Data Availability

All data produced in the present study are available upon reasonable request to the authors

## Acknowledgements

This work was funded by the Government of Alberta. The authors wish to thank Dr. Norma Ruecker and Dr. Rhonda Clark for help with sampling and logistics. This work would not have been possible without collaboration from provincial and municipal leaders and WWTP operators in Calgary, Edmonton, Fort McMurray, Grande Prairie, Cold Lake, Edson, Lacombe, Red Deer, Banff, Canmore, Drumheller, Strathmore, Okotoks, High River, Brooks, Medicine Hat, Taber and Lethbridge.

---

**Table S1.** Chronology of detection of the Omicron variant in Alberta community wastewater

| WWTP or pump station | Date of 1<sup>st</sup> Omicron detection | ≥ 50% Omicron detected | ≥ 85% Omicron detection | ≥ 99% Omicron detection |
| --- | --- | --- | --- | --- |
| Airdrie | Dec 16, 2021 | – | Dec 16, 2021 | Jan 5, 2022 |
| Banff | Dec 12, 2021 | Dec 14, 2021 | Dec 22, 2021 | Dec 27, 2021 |
| Brooks | Dec 29, 2021 | Dec 29, 2021 | Jan 5, 2022 | Jan 5, 2022 |
| Calgary-1<sup>a</sup> | Dec 1, 2021 | Dec 15, 2021 | Dec 26, 2021 | Dec 29, 2021 |
| Calgary-1<sup>b</sup> | Nov 30, 2021 | Dec 20, 2021 | Dec 28, 2021 | Jan 3, 2022 |
| Calgary-2 | Dec 7, 2021 | Dec 19, 2021 | Dec 26, 2021 | Jan 4, 2022 |
| Calgary-3 | Dec 7, 2021 | Dec 19, 2021 | Dec 26, 2021 | Jan 12, 2022 |
| Canmore | Dec 13, 2021 | Dec 20, 2021 | Dec 29, 2021 | – |
| Cold Lake | Dec 16, 2021 | Dec 29, 2021 | Jan 5, 2022 | Jan 11, 2022 |
| Drumheller | Dec 20, 2021 | Dec 20, 2021 | Dec 29, 2021 | Jan 17, 2022 |
| Edmonton-1 | Dec 10, 2021 | Dec 22, 2021 | Dec 28, 2021 | Jan 17, 2022 |
| Fort McMurray<sup>a</sup> | Dec 6, 2021 | Dec 15, 2021 | Dec 22, 2021 | Dec 22, 2021 |
| Fort McMurray<sup>b</sup> | Dec 1, 2021 | Dec 20, 2021 | Dec 22, 2021 | Jan 5, 2022 |
| Edmonton-2 | Dec 8, 2021 | Dec 22, 2021 | Dec 29, 2021 | Jan 14, 2022 |
| Grande Prairie | Dec 20, 2021 | Dec 20, 2021 | – | – |
| High River | Dec 20, 2021 | Dec 22, 2021 | Dec 29, 2021 | Dec 30, 2021 |
| Lacombe | Dec 16, 2021 | NA | Jan 5, 2022 | – |
| Lethbridge<sup>b</sup> | Dec 14, 2021 | Dec 21, 2021 | Dec 29, 2021 | Jan 10, 2022 |
| Lethbridge<sup>a</sup> | Dec 19, 2021 | – | Dec 28, 2021 | Jan 5, 2022 |
| Medicine Hat | Dec 16, 2021 | Dec 23, 2021 | Dec 30, 2021 | Jan 4, 2022 |
| Okotoks | Dec 13, 2021 | Dec 20, 2021 | Dec 20, 2021 | Jan 6, 2022 |
| Red Deer | Nov 29, 2021 | Dec 23, 2021 | Jan 4, 2022 | Jan 17, 2022 |
| Strathmore | Dec 15, 2021 | Dec 20, 2021 | Jan 5, 2022 | Jan 19, 2022 |
| Taber | Dec 29, 2021 | NA | Dec 29, 2021 | Jan 5, 2022 |

<sup>a</sup> wastewater samples processed using the affinity column method
<sup>b</sup> wastewater samples processed using the ultrafiltration method

**Figure S1.**
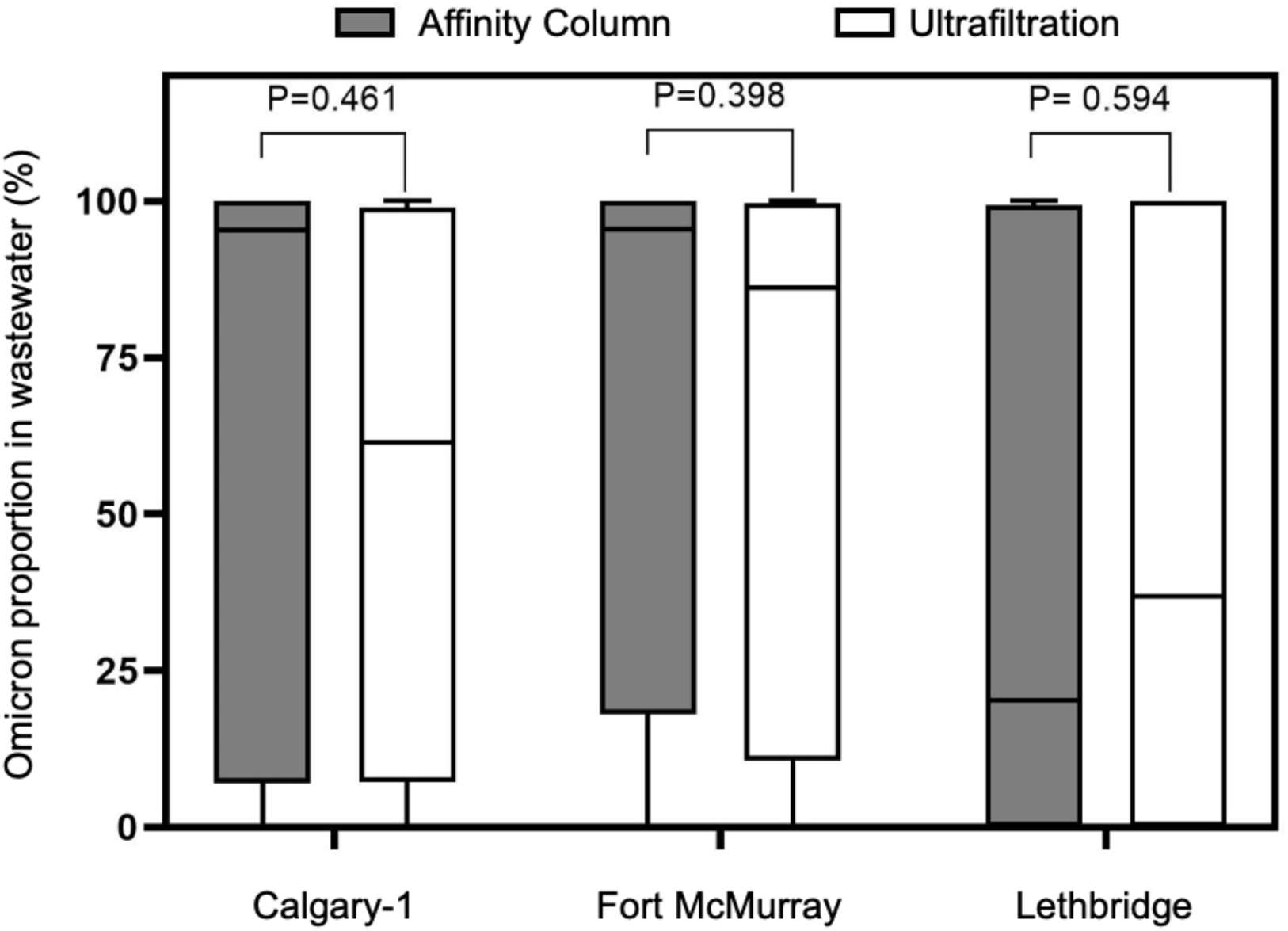
Wastewater sample processing using affinity columns and ultrafiltration was compared by testing samples from WWTPs in Calgary-1 (n=14), Fort McMurray (n=18) and Lethbridge (n=15) using both methods. The proportion of Omicron obtained following either processing method was compared using Mann-Whitney tests revealing no significant difference in median values between the two sample processing methods. Median and interquartile ranges are indicated as the middle, top, and bottom lines of each box. Ends of the whiskers mark the lowest and highest ratios determined in each sample series.

**Figure S2.**
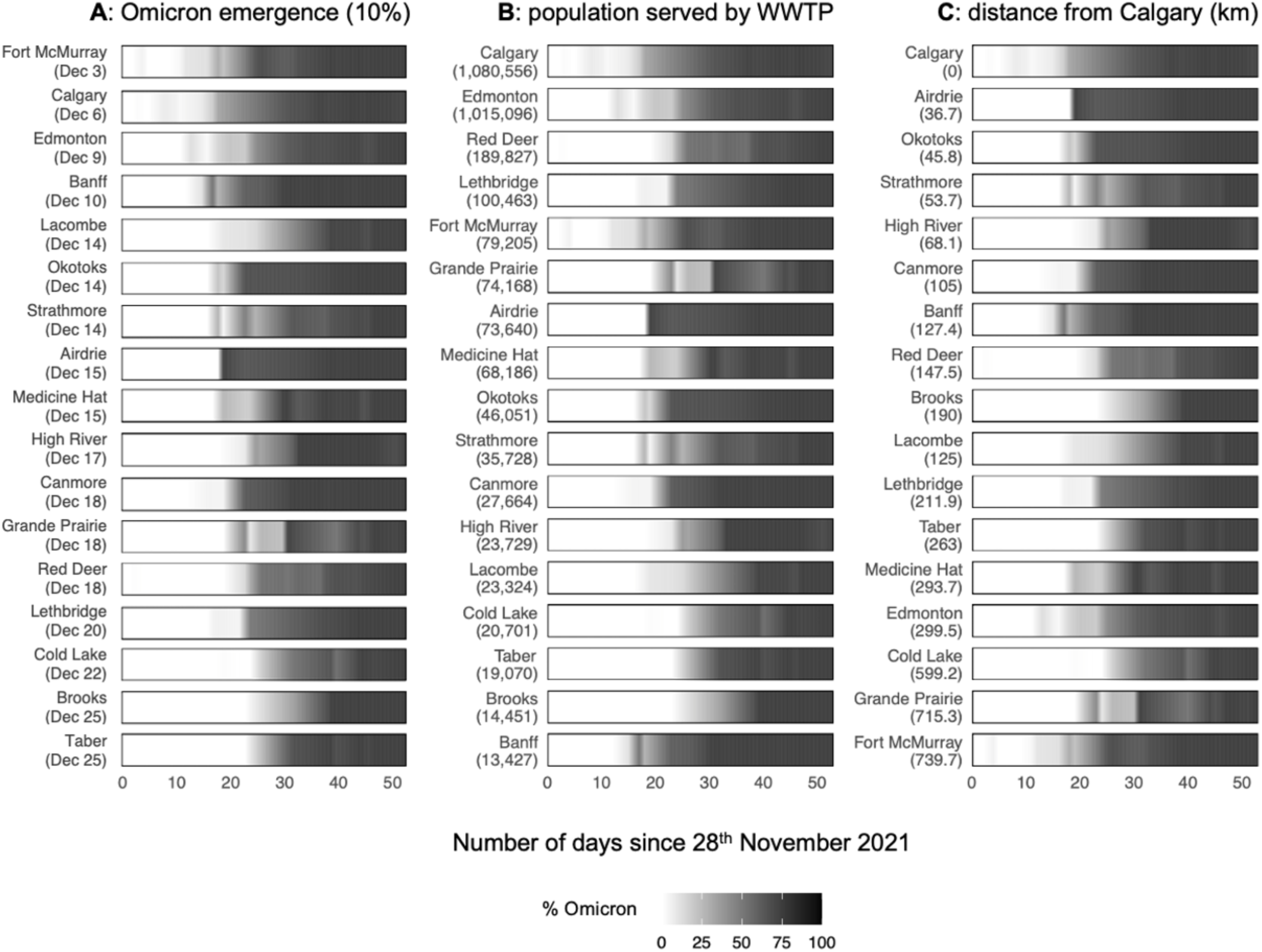
Proportion of Omicron in Alberta municipalities plotted as a function of (A) the timing of its emergence, (B) the population served by the municipal WWTP, and (C) the distance of the municipality from Calgary. Timing in panel A corresponds to the midpoint between sampling dates with values below and above 10%. Panels B and C highlight communities that don’t follow the general trends of Omicron emerging earlier in larger municipalities (e.g., Banff has the smallest resident population but increases early), and emerging later in locations farther away Calgary’s major international airport (e.g., Fort McMurray is the farthest away from Calgary but increases early), respectively. Edmonton and Calgary results are limited to the largest WWTPs in the two cities (i.e., Edmonton-1 and Calgary-1 from Figure 1).

## Literature Cited

Acosta N, Bautista M, Hollman J, McCalder J, Buchner Beaudet A, Man L, Waddell BJ, Chen J, Li C, Kuzma D, Bhatnagar S, Leal J, Meddings J, Hu J, Cabaj J, Ruecker NJ, Naugler C, Pillai DR, Achari G, Ryan CM, Conly JM, Frankowski K, Hubert CRJ, Parkins MD (2021a) Wastewater monitoring of SARS-CoV-2 from acute care hospitals identifies nosocomial transmission and outbreaks. Water Research 201: 117369. https://doi.org/10.1016/j.watres.2021.117369

Acosta N, Bautista MA, Waddell BJ, McCalder J, Buchner Beaudet A, Man L, Pradhan P, Sedaghat N, Papparis C, Bacanu A, Hollman J, Krusina A, Southern D, Williamson T, Li C, Bhatnagar S, Murphy S, Chen J, Kuzma D, Meddings J, Hu J, Cabaj JL, Conly JM, Ruecker NJ, Achari G, Ryan CM, Frankowski K, Hubert CRJ, Parkins MD. (2021b) Longitudinal SARS-CoV-2 RNA Wastewater Monitoring Across a Range of Scales Correlates with Total and Regional COVID-19 Burden in a Well-Defined Urban Population. Submitted to medRxiv doi: https://www.medrxiv.org/content/10.1101/2021.11.19.21266588v1

Ahmed W, Bertsch PM, Bivins A, Bibby K, Farkas K, Gathercole A, Haramoto E, Gyawali P, Korajkic A, McMinn BR, Mueller JF, Simpson SL, Smith WJM, Symonds EM, Thomas KV, Verhagen R, Kitajima M (2020) Comparison of virus concentration methods for the RT-qPCR based recovery of murine hepatitis virus, a surrogate for SARS-CoV-2 from untreated wastewater. Science of the Total Environment 739: 139960.

AHS (2022) Assessment and Testing COVID-19. Alberta Health Services https://www.albertahealthservices.ca/topics/Page17058.aspx. Accessed February, 2022.

Alberta Health (2022) Covid-19 Alberta Statistics. Government of Alberta https://www.alberta.ca/stats/covid-19-alberta-statistics.htm#variants-of-concern. Accessed February, 2022.

Banff (2022) “Learn about Banff” website: https://banff.ca/252/Learn-About-Banff. Town of Banff. Accessed February, 2022.

D’Aoust, PM, Graber, TE, Mercier, E, Montpetit, D, Alexandrov, I, Neault, N, Baig, AT, Mayne, J, Zhang, X, Alain, T, Servos, MR, Srikanthan, N, MacKenzie, M, Figeys, D, Manuel, D, Jüni, P, MacKenzie, AE, Delatolla, R (2021) Catching a resurgence: Increase in SARS-CoV-2 viral RNA identified in wastewater 48 h before COVID-19 clinical tests and 96 h before hospitalizations. Science of the Total Environment. 770: 145319.

WHO (2021) Tracking SARS-CoV-2 variants. World Health Organization. https://www.who.int/en/activities/tracking-SARS-CoV-2-variants/. Accessed February, 2022.

Cevik M, Tate M, Lloyd O, Maraolo AE, Schafers J, Ho A (2021) SARS-CoV-2, SARS-CoV, and MERS-CoV viral load dynamis, duration of viral shedding, and infectiousness: a systematic review and meta-analysis. The Lancet Microbe 2: E13–E22.

Chavarria-Miró G, Anfruns-Estrada E, Martínez-Velázquez A, Vázquez-Portero M, Guix S, Paraira M, Galofré B, Sánchez G, Pintó RM, Bosch A (2021) Time evolution of severe acute respiratory syndrome coronavirus 2 (SARS-CoV-2) in wastewater during the first pandemic wave of covid-19 in the metropolitan area of Barcelona, Spain. Applied & Environmental Microbiology 87: e02750–20.

Elbe S, Buckland-Merrett G (2017) Data, disease and diplomacy: GISAID’s innovative contribution to global health. Global Challenges 1: 33–46.

Graber TE, Mercier É, Bhatnagar K, Fuzzen M, D’Aoust PM, Hoang HD, Tian X, Towhid ST, Plaza-Diaz J, Eid W, Alain R, Butler A, Goodridge L, Servos M, Delatolla R. (2021). Near real-time determination of B.1.1.7 in proportion to total SARS-CoV-2 viral load in wastewater using an allele-specific primer extension PCR strategy. Water Research 205: 117681. https://doi.org/10.1016/j.watres.2021.117681

Green MA, García-Fiñana M, Barr B, Burnside F, Cheyne CP, Hughes D, Ashton M, Sheard S, Buchan IE (2021) Evaluating social and spatial inequalities of large scale rapid lateral flow SARS-CoV-2 antigen testing in COVID-19 management: An observational study of Liverpool, UK (November 2020 to January 2021). The Lancet Regional Health Europe 6: 100107.

Lee WL, Imakaev M, Armas F, McElroy KA, Gu X, Duvallet C, Chandra F, Chen H, Leifels M, Mendola S, Floyd-O’Sullivan R, Powell MM, Wilson ST, Berge KLJ, Lim CYJ, Wu F, Xiao A, Moniz K, Ghaeli N, Matus M, Thompson J, Alm EJ (2021) Quantitative SARS-CoV-2 Alpha Variant B.1.1.7 Tracking in Wastewater by Allele-Specific RT-qPCR. Environmental Science & Technology Letters https://doi.org/10.1021/acs.estlett.1c00375

Lin X, Glier M, Kuchinski K, Ross-Van Mierlo T, McVea D, Tyson JR, Prystajecky N, Ziels RM (2021) Assessing Multiplex Tiling PCR Sequencing Approaches for Detecting Genomic Variants of SARS-CoV-2 in Municipal Wastewater. mSystems 6: e01068–21.

Medema G, Heijnen L, Elsinga G, Italiaander R, Brouwer A (2020) Presence of SARS-Coronavirus-2 RNA in sewage and correlation with reported COVID-19 prevalence in the early stage of the epidemic in the Netherlands. Environmental Science and Technology Letters 7: 511–516.

Naughton C, Román F, Alvarado AG, Tariqi AQ, Deeming M, Bibby K, Bivins A, Rose J, Medema G, Ahmed W, Katsivelis P, Allan V, Sinclair R, Zhang Y, Kinyua M (2021) Show us the Data: Global COVID-19 Wastewater Monitoring Efforts, Equity, and Gaps. Published in medRxiv 17 March 2021. DOI:10.1101/2021.03.14.21253564.

Nemudryi A, Nemudraia A, Wiegand T, Surya K, Buyukyoruk M, Cicha C, Vanderwood KK, Wilkinson R, Wiedenheft B (2020) Temporal detection and phylogenetic assessment of SARS-CoV-2 in municipal wastewater. Cell Reports Medicine 1: 100098.

Pecson BM, Darby E, Haas CN, Amha YM, Bartolo M, Danielson R, Dearborn Y, Di Giovanni G, Ferguson C, Fevig S, Gaddis E, Gray D, Lukasik G, Mull B, Olivas L, Olivieri A, Qucand Y, SARS-CoV-2 Interlaboratory Consortium (2021) Reproducibility and sensitivity of 36 methods toquantify the SARS-CoV-2 genetic signal in rawwastewater: findings from an interlaboratorymethods evaluation in the U.S. Environmental Science: Water Research and Technology 7: 504.

Pérez-Cataluña A, Chiner-Oms A, Cuevas-Ferrando E, Díaz-Reolid A, Falcó I, Randazzo W, Girón-Guzmán I, Allende A, Bracho MA, Comas I, Sánchez G (2022) Spatial and temporal distribution of SARS-CoV-2 diversity circulating in wastewater. Water Research 211: 118007.

Peterson SW, Lidder R, Daigle J, Wonitowy Q, Dueck C, Nagasawa A, Mulvey MR, Mangat CS (2022) RT-qPCR detection of SARS-CoV-2 mutations S 69-70 del, S N501Y and N D3L associated with variants of concern in Canadian wastewater samples. Science of the Total Environment 810: 151283. https://doi.org/10.1016/j.scitotenv.2021.151283

Qiu Y, Yu J, Pabbaraju K, Lee BE, Gao T, Ashbolt NJ, Hrudey SE, Diggle M, Tipples G, Maal-Bared R, Pang X (2022) Validating and optimizing the method for molecular detection and quantification of SARS-CoV-2 in wastewater. Science of the Total Environment 812: 151434. https://doi.org/10.1016/j.scitotenv.2021.151434

Rios G, Lacoux C, Leclercq V, Diamant A, Lebrigand K, Lazuka A, Soyeux E, Lacroix S, Fassy J, Couesnon A, Thiery R, Mari B, Pradier C, Waldmann R, Barbry P (2021) Monitoring SARS-CoV-2 variants alternations in Nice neighborhoods by wastewater nanopore sequencing. Lancet Regional Health – Europe 10: 100202.

Randazzo W, Truchado P, Ferranfo EC, Simon P, Allende A, Sanchez G (2020) SARS-CoV-2 RNA titers in wastewater anticipated COVID-19 occurrence in a low prevalence area. Water Research 181: 115942.

Rothman JA, Loveless TB, Kapcia J, Adams ED, Steele JA, Zimmer-Faust AG, Langlois K, Wanless D, Griffith M, Mao L, Chokry J, Griffith JF, Whiteson KL (2021) RNA viromics of Southern California wastewater and detection of SARS-CoV-2 single nucleotide variants. Applied and Environmental Microbiology 01448–21

Whitney ON et al (2021) Sewage, Salt, Silica, and SARS-CoV-2 (4S): An Economical Kit-Free Method for Direct Capture of SARS-CoV-2 RNA from Wastewater. Environmental Science & Technology. 55: 4880–4888.

Williams GH, Llewelyn A, Brandao R, Chowdhary K, Hardisty KM, Loddo M (2021) SARS-CoV-2 testing and sequencing for international arrivals reveals significant cross border transmission of high risk variants into the United Kingdom. eClinicalMedicine Lancet 38: 101021.

YEG (2022) Edmonton International Airport. https://flyeia.com. Accessed February, 2022.

YMM (2022) Fort McMurray Airport Authority Annual Report. https://www.flyymm.com/publications. Accessed February, 2022.

YYC (2022) Calgary International Airport. www.yyc.com. Accessed February, 2022.

Yuan C, Wang H, Li K, Tang A, Dai y, Wu B, Zhang H, Chen J, Liu J, Wu W, Gu S, Wang H, Xu H, Wu M, Yu M, Wang Y, Yu X, He J, Liu S, Zhang Y, Tong Z, Yan J (2021) SARS-CoV-2 viral shedding characteristics and potential evidence for the priority for faecal specimen testing in diagnosis. PLoS One 16: e0247367.

